# Burden of health morbidities and associated health care costs in the Australian Genetics of Depression Study using the medication-based Rx-Risk Comorbidity Index

**DOI:** 10.64898/2026.05.15.26353340

**Authors:** Penelope A Lind, Ian B. Hickie, Enda M Byrne, Nicholas G. Martin, Sarah E Medland

## Abstract

Depression is accompanied by considerable comorbidity and excess mortality. We examined multimorbidity data using the validated pharmacy-based Rx-Risk Comorbidity Index and examined healthcare costs associated with chronic illness burden in the Australian Genetics of Depression Study (AGDS). Australian Pharmaceutical Benefits Scheme (PBS) record linkage for 15,890 AGDS participants was available from 01/07/2013-31/12/2017. Forty-six health morbidities were inferred by mapping the prescription data using Anatomical Therapeutic Chemical Classification System codes and PBS Item Codes. Morbidity prevalence rates were then compared with an unselected 10% Australian representative population sample (10PCT) with PBS claims data available from 01/07/2010-31/12/2014. The average number of inferred comorbidities was higher among AGDS participants (4.6 ± 2.9) than 10PCT individuals (3.0 ± 3.0). Excluding depression, 89.1% of AGDS participants had one or more inferred comorbidity, most commonly pain (51.0%), inflammation/pain (40.3%), and anxiety (32.3%). In the AGDS, the number of comorbidities was higher among women compared to men and positively correlated with participant age, BMI, number of depressive episodes experienced, and annual health care costs. Compared to participants with no inferred comorbidities, the median annual health care costs were ∼65% higher among those with 2-3 comorbidities. This study highlights the patterns of health morbidities experienced by individuals living with depression and shows that this chronic disease burden is significantly associated with increased health costs to the individual and the health system.

## 1. Introduction

Major depressive disorder (MDD) is a common mental health disorder accompanied by considerable comorbidity [1, 2] and excess mortality [1, 3]. Individuals living with MDD face an elevated, and sometimes premature, burden of poor physical health outcomes, including increased risk of cancer [4], a 3-fold increased risk for cardiovascular disease (CVD) which manifests 7 years earlier [5], a 2-fold increased risk for type 2 diabetes mellitus, 3-fold increased risk for stroke, and a 5-fold increased risk of myocardial infarction [6]. Of note, depression has been found to be a significant risk factor in the development of coronary heart disease (CHD) [7] and cardiovascular morbidity and mortality is significantly higher in individuals who experience symptoms of depression as well as individuals diagnosed with MDD [5].

Critically, individuals with lived experience of moderate to severe MDD die 10 to 15 years earlier than the general population due to both natural and unnatural causes [8, 9]. Furthermore, a recent study of the gap in life expectancy in psychiatric patients compared with the general population in Western Australia found that almost 75% of excess deaths in people living with depression were due to physical health conditions, with cardiovascular disease and cancer accounting for approximately 45% of excess deaths [9]. Aetiological factors involved may include adverse effects of medication, lifestyle risk factors such as substance use [5, 10], and poorer access to and quality of physical health care [11, 12].

In this study, we sought to examine the relationship between depression and its comorbidities in 15,890 participants from the Australian Genetics of Depression Study (AGDS) [13], a large study of the genetic causes of depression and treatment response. Since the AGDS cohort lacks clinical diagnosis data, the Rx-Risk Comorbidity Index [14] was used to identify 46 chronic conditions from 4.5 years of pharmaceutical claims data. The large sample size of the AGDS, the rich phenotypic data collected, and the enrichment of patients living with recurrent severe depression, make this a unique cohort to examine comorbidity patterns and risk factors for chronic disease burden in an Australian cohort of individuals living with depression, along with the impact of this burden on healthcare costs.

## 2. Methods

### 2.1 Participants

#### 2.1.1 Australian Genetics of Depression Study (AGDS)

Participants were drawn from the Australian Genetics of Depression Study[13], a nationwide cohort of 20,689 adults living with depression (75% female; aged 43±15 years [range 18–90]) recruited between 2017 and 2020. The AGDS cohort is enriched for recurrent severe depression. Participants self-reported an average of 7.95 episodes of depression in their lifetime, and 5,990 participants experienced 10 or more episodes of depression. Approximately 75% of participants consented to record linkage of their Pharmaceutical Benefits Schedule (PBS) data for the previous 4.5 years and provided their name, address, and Medicare card number.

AGDS consent forms were sent to *Services Australia* in November 2019 to request approval to link the participant’s electronic PBS and Medicare Benefits Schedule (MBS) records. *Services Australia* supplied the MBS and PBS data to QIMR Berghofer. The PBS research database contains pharmacy dispensing records for most Australians for the previous 4.5 calendar years. Those records include all medications listed on the PBS Schedule that are subsidized by the Australian Government for eligible Medicare card holders. PBS records provide PBS Item Codes and Anatomical Therapeutic Chemical (ATC) Classification System codes for each dispensed medication as well as costs to the Medicare health system (Medicare benefits paid) and out-of-pocket costs for the individual. The MBS records include data for each health service accessed, including all consultations and diagnostic procedures and tests outside the hospital setting, as well as cost data for each health service; these allow us to calculate the total annual out-of-pocket costs to the individual and Medicare benefits paid. The PBS and MBS records do not contain indication or diagnosis information. Approximately 75% of all prescriptions dispensed in Australia are recorded in the PBS database [15].

The window for data extraction was from 01/07/2013 and 31/12/2017 (4 years and 6 months). Data linkage was only available for participants who consented and provided a valid Medicare number and address and had a history of at least one dispensed medication or accessed medical service in that window. Overall, data linkage was obtained for 15,890 participants of whom 13,570 participants (85.4%) met *Diagnostic and Statistical Manual of Mental Disorders* [5th ed.; DSM-5; 16] criteria for lifetime MDD, 1197 screened positive for depression in the previous two weeks, and 311 did not meet criteria for DSM-5 MDD or screen positive. The PBS and MBS data were linked to the AGDS baseline survey data.

#### 2.1.2 10% Sample of the Australian PBS Claims Database (10PCT)

We analysed data from a publicly available deidentified and unselected 10% population sample of the Australian PBS claims database published by *Services Australia* on 1 August 2016 [17]. The PBS claims represent all dispensed prescriptions from 2003 to 2014 for a randomly selected 10% sample of the Australian population (N=2,985,512). The sample was deidentified and included sex, year of birth, and the State of residence. Data provided for each dispensed medication included the dates of prescribing and medication supply, the medication name, PBS Item Code, and ATC code. For the current study, PBS records in the most recent 4.5-year window (01/07/2010 and 31/12/2014) and participants aged between 18 and 90 years old (n=1,636,478; 52.6% female) were included in analyses, to match the AGDS 4.5-year window of PBS record linkage and participant age range.

### 2.2 Mapping of Comorbidity Categories from Dispensed Prescription Data

The Rx-Risk Comorbidity Index model developed by Pratt et al. [14] was used to map dispensed medication data to 46 comorbidity categories using ATC codes and PBS Item Codes in both the AGDS and 10PCT samples. An unweighted Rx-Risk Comorbidity Index score (referred to as the Rx-Risk score) was calculated as the number of comorbidity categories that an individual was treated for with one or more dispensed medications indicative of that condition. It should be noted that the 46 comorbidities mapped using this model are only inferred by prescription data and do not indicate confirmed diagnoses. However, for simplicity within this study, the language used with respect to these comorbidities does not always indicate that they are inferred only.

### 2.3 Health Care Costs

A secondary analysis of the impact of multimorbidity on health care costs was conducted in 15,022 participants (11,186 females and 3,836 males) who had PBS prescription dispensing data and/or MBS medical service use records for the full 4.5-year window (01/07/2013 to 31/12/2017). Total PBS out-of-pocket costs paid by the participant and the Medicare benefits paid (Medicare subsidy) for dispensed prescriptions each year were calculated using ‘Patient Contribution’ and ‘Net Benefit’ variables respectively. Total annual MBS out-of-pocket costs paid by the participant and the Medicare benefits paid (Medicare subsidy) for services each year were calculated using ‘Patient Out of Pocket’ and ‘Benefit Paid’ variables respectively. Annual total PBS and MBS costs were then calculated by dividing the total costs by the number of years (1 to 4.5) for which the PBS and MBS data were available respectively. The annual PBS and MBS costs were then summed to estimate the total annual health care out-of-pocket costs to the participant and costs to the Medicare health system (Medicare benefits paid).

### 2.4 Ethics

Ethics approval for all aspects of the project was obtained from the QIMR Berghofer Human Research Ethics Committee (P2118). In addition, the Study mail-out and linkage of participant PBS and MBS data were also approved by the *Services Australia* External Request Evaluation Committee (EREC reference number MI3967). The Helsinki Declaration, as well as applicable institutional and governmental regulations concerning the ethical use of human volunteers were followed during all the phases of this research.

### 2.5 Statistical analyses

Rx-Risk score calculation and basic statistical analysis, including descriptive analyses, were conducted using IBM SPSS Statistics for Windows, version 23.0 (IBM Corp., Armonk, NY, USA). Logistic regression was used to assess sex differences in the prevalence of each Rx-Risk comorbidity; each result is presented as an odds ratio (OR) and 95 % confidence interval (95%CI). Mann-Whitney U tests were used to compare the number of inferred morbidities (the Rx-Risk score) according to the sample (AGDS vs 10PCT), sex within each sample, as well as age, depression severity measures, and body mass index (BMI) ranges in the AGDS sample.

In the latter analyses, participant age ranges were grouped in decades (<20, 20-29, 30-39, 40-49, 50-59, 60-69, 70+ years) while BMI ranges (Underweight (BMI < 18.5), normal weight [18.5 ≤ BMI > 25], overweight [25 ≤ BMI > 30], obesity class I [30 ≤ BMI > 35], obesity class II [35 ≤ BMI > 40], obesity class III [BMI ≥ 40]) were aligned to the World Health Organization *Nutritional Status* categories [18].

The self-reported number of depressive episodes experienced by participants ranging from 1 (n = 579, 3.6%) to 13 or more (n = 5147, 32.4%) was grouped as follows: 1, 2-3, 4-5, 6-7, 8-9, 10-12, 13+ episodes.

Finally, Spearman’s rank correlations (with bias-corrected and accelerated [BCa] bootstrap 95% confidence intervals) were computed to examine the relationship between annual out-of-pocket costs for participants and the Medicare benefits paid (the Medicare subsidy) and the number of inferred morbidities (after the depression comorbidity category was excluded) among AGDS participants. All costs are presented in Australian dollars at the time of dispensing (2013-2017).

## 3. Results

Pharmaceutical claims data were available for a total of 15,890 AGDS participants and over 1.6 million individuals from the 10% Sample of the Australian PBS Claims Database. The mean (SD) participant age at the time of the AGDS survey was 44.3 (15.2) years, the median 44 years and the range 18–90 years (Table 1). AGDS participants were on average younger than those in the 10PCT sample who were aged 48.4 (18.5) years, median 47 years and range 18–90 years. Supplemental Figure S1 shows the distribution of ages in the AGDS and 10PCT samples.

**Table 1.** Distribution of participant age and Rx-Risk scores by sample and sex.

| Sample and Variable | AGDS |  | 10PCT |  |
| --- | --- | --- | --- | --- |
|  | Male | Female | Male | Female |
| Participants, n (%) | 4196 (26.4) | 11694 (73.6) | 775683 (47.4) | 860795 (52.6) |
| Participant age in years, % |  |  |  |  |
| Mean (SD) | 48.7 (15.0) | 42.7 (15.0) | 48.6 (18.3) | 48.3 (18.6) |
| Range (minimum – maximum)<sup>b</sup> | 18 – 89 | 18 – 90 | 18 – 90 | 18 – 90 |
| 18-19 | 0.5 | 1.8 | 3.1 | 3.0 |
| 20-29 | 12.5 | 23.1 | 15.6 | 16.5 |
| 30-39 | 17.6 | 21.4 | 16.5 | 17.3 |
| 40-49 | 18.4 | 18.9 | 17.4 | 17.4 |
| 50-59 | 23.7 | 18.5 | 17.3 | 16.6 |
| 60-69 | 19.9 | 12.6 | 15.0 | 14.1 |
| 70+ | 7.4 | 3.7 | 15.1 | 15.3 |
| Adjusted Rx-Risk Score<sup>a</sup>, % |  |  |  |  |
| Mean (SD) | 3.9 (3.0) | 3.6 (2.8) | 2.7 (2.8) | 2.8 (2.9) |
| Range (minimum – maximum) | 0 – 16 | 0 – 17 | 0 – 20 | 0 – 20 |
| 0<sup>c</sup> | 11.4 | 10.7 | 20.1 | 21.6 |
| 1 | 13.7 | 14.2 | 23.3 | 21.1 |
| 2-3 | 27.3 | 30.3 | 27.0 | 26.5 |
| 4-6 | 27.7 | 29.4 | 18.4 | 18.8 |
| 7-10 | 16.6 | 13.2 | 9.3 | 9.9 |
| 11-13 | 3.0 | 2.0 | 1.8 | 2.0 |
| 15-18 | 0.2 | 0.1 | 0.11 | 0.14 |
| 19+ | 0 | 0 | 0.001 | 0.002 |

Abbreviations: 10PCT, 10% Sample of the Australian PBS Claims Database; AGDS, Australian Genetics of Depression Study; SD, standard deviation.
<sup>a</sup>Defined as the number of different comorbidity categories after excluding depression mapped by prescriptions dispensed between 01/07/2013 and 31/12/2017, possible range 0-45.
<sup>b</sup>Age ranges matched.
<sup>c</sup>These individuals had not been prescribed any of the medications used in the Rx-Risk algorithm.

### Burden of comorbidities

The total number of comorbidities (Rx-Risk score) inferred from the AGDS prescription data ranged from 0 to 17 among male participants (*x̅* = 4.79, *s* = 3.10, median = 4) and 0 to 18 among female participants (*x̅* = 4.55, *s* = 2.84, median = 4). The overall burden of comorbidities in the unselected population sample (10PCT) was lower, where the average number of comorbidities among males and females were 2.9 ± 2.9 and 3.1 ± 3.0, respectively (see Supplemental Figure S2).

Overall, 87.2% of AGDS participants and 60.0% of 10PCT individuals had two or more inferred comorbidities (referred to as multimorbidity); the rate of multimorbidity reduced to 75.0% among AGDS participants and 56.9% among individuals in the 10PCT after excluding the depression comorbidity which had a prevalence rate of 90.7% and 23.1% in the AGDS and 10PCT cohorts respectively (Supplemental Table S1).

After excluding the depression morbidity category, 20.9% of 10PCT individuals and 10.9% of AGDS participants had no inferred comorbidities and the average number of comorbidities in the 10PCT and AGDS samples was reduced to 2.8 ± 2.8 and 3.7 ± 2.9, respectively (Table 1). In addition, after excluding depression, individuals in the 10PCT sample were significantly more likely to have no inferred comorbidities (Odds ratio [OR]= 2.17, 95% confidence interval [95%CI]: 2.07-2.28, p < 0.001) and 2.3-fold less likely (Odds ratio [OR]= 0.44, 95%CI: 0.42-0.46, p < 0.001) to experience multimorbidity than AGDS participants.

The mean rank of Rx-Risk scores in AGDS was significantly lower among females than males although the effect size was small, *z* =-3.113, *p* = 0.0019, *r* = −0.025. In addition, Jonckheere-Terpstra tests show that as participant age (*z* = 41.535, *p* < 0.001, *r* = 0.35), obesity levels (*z* = 33.779, *p* < 0.001, *r* = 0.29) and self-reported number of depressive episodes (*z* = 21.300, *p* < 0.001, *r* = 0.18) increased, the median Rx-Risk score significantly increased. This pattern of association was also observed in males and females separately (Supplemental Figures S3-5). Similarly, increases in participant age in the 10PCT sample were also associated with significantly higher Rx-Risk scores (*z* = 700.549, *p* < 0.001, *r* = 0.55) but the mean rank of Rx-Risk scores was significantly higher among females than males, *z* = 24.215, *p* < 0.001, r = 0.019.

### Prevalence of comorbidities in AGDS and 10PCT

Figure 1 illustrates the inferred prevalence of each comorbidity category in the AGDS and 10PCT samples by gender with the prevalence rates given in Supplemental Table S1. No AGDS participant or 10PCT individual was dispensed medications for hyperkalaemia in the 4.5-year window and AGDS participants also did not receive medications for malnutrition. As expected, depression was the most common inferred morbidity among AGDS participants (90.7%) while the prevalence was 23.1% in the 10PCT sample. Compared to the 10PCT sample, the prevalences of other mental health conditions (alcohol dependency, anxiety, bipolar disorder, psychotic illness) were also higher among AGDS participants.

**Figure 1.**
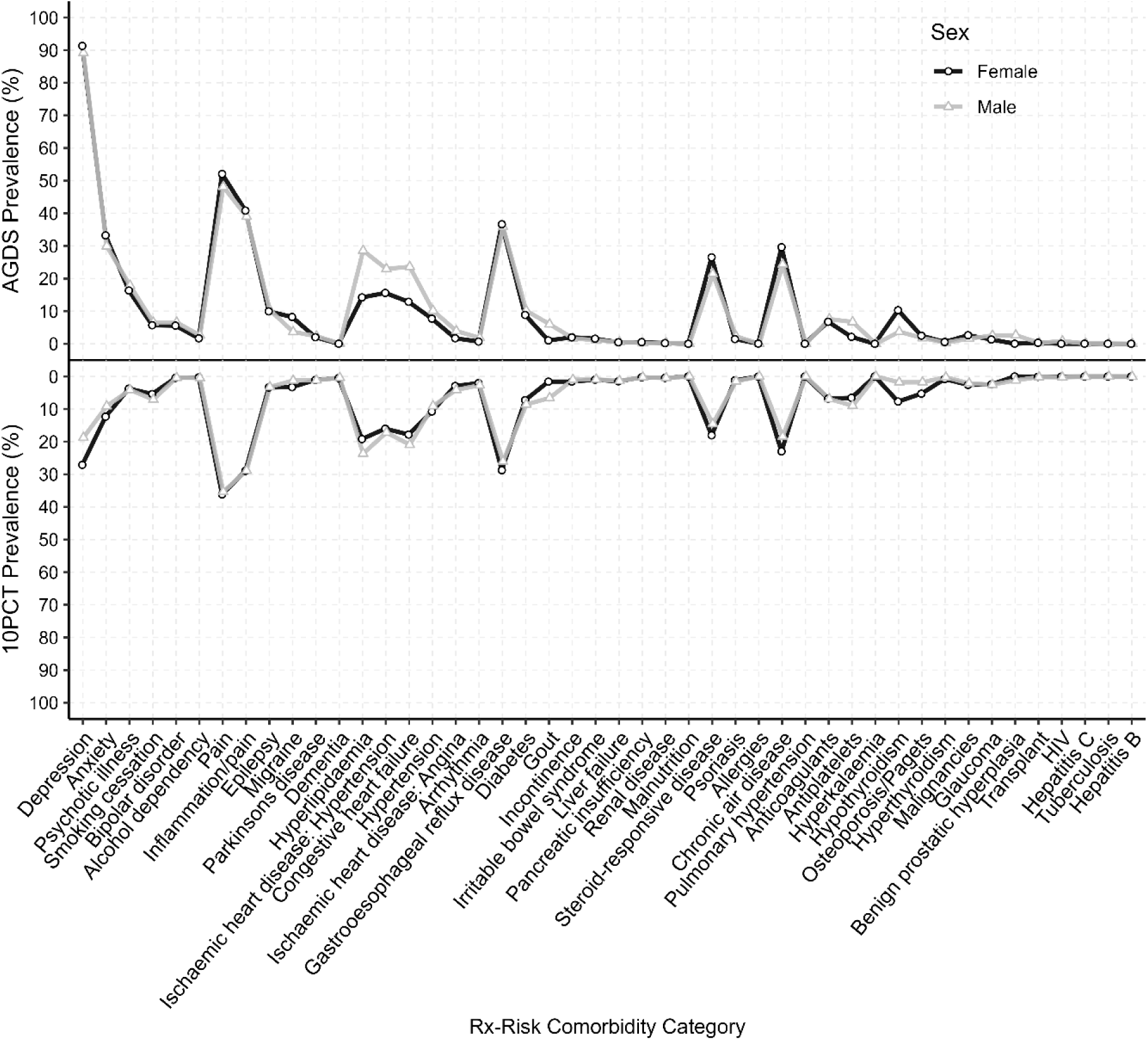
The prevalence of each inferred Rx-Risk comorbidity category in male and female participants from the Australian Genetics of Depression Study (AGDS; top panel) and 10% Sample of the Australian PBS Claims Database (10PCT; bottom panel).

Excluding depression, 42.2% of AGDS participants had one or more mental health condition compared to only 13.3% of individuals in the 10PCT sample. However, the prevalence of dementia was 8-fold higher in the 10PCT sample, possibly due to higher proportion of individuals aged 70 or over in the 10PCT sample (15.2%) compared to AGDS (4.7%). AGDS participants were also more likely than the general population (85.7% vs 76.8%) to have received medications for one or more physical comorbidity.

The impact of sex and sample (after adjusting for participant age) on the odds of having an inferred comorbidity is summarised in Table 2. For each of the comorbidities observed in both male and female AGDS participants, the prevalence was highest for males versus females for Human Immunodeficiency Virus (HIV; OR=98.1, 95%CI:21.1-1747.4, *p* < 0.001), gout (OR=4.84, 95%CI:3.88-6.07, *p* < 0.001), hepatitis C (OR=3.91, 95%CI:0.94-19.43, *p* < 0.001) and antiplatelet use (OR=2.26, 95%CI:1.89-2.71, *p* < 0.001). Compared to female AGDS participants, males were much less likely to have been dispensed medications for hypothyroidism (OR=0.27, 95%CI:0.22-0.32, *p* < 0.001) and Osteoporosis/Paget’s disease (OR=0.40, 95%CI:0.31-0.53, *p* < 0.001).

**Table 2.** Adjusted odds ratio of each comorbidity category associated with sample and sex.

| Reference Group: | AGDS Females | AGDS Males | AGDS Females | 10PCT Females |
| --- | --- | --- | --- | --- |
| Comparison Group: | AGDS Males | 10PCT Males | 10PCT Females | 10PCT Males |
| Alcohol dependency | 1.50 (1.17-1.90) | **0.19 (0.16-0.23)** | **0.17 (0.15-0.20)** | **1.72 (1.63-1.81)** |
| Anticoagulants | 0.87 (0.76-1.00) | **0.73 (0.65-0.82)** | **0.65 (0.60-0.70)** | 1.02 (1.01-1.03) |
| Antiplatelets | **2.26 (1.89-2.71)** | 1.02 (0.90-1.16) | **1.38 (1.22-1.58)** | **1.56 (1.54-1.58)** |
| Anxiety | **0.85 (0.79-0.92)** | **0.23 (0.22-0.25)** | **0.25 (0.24-0.26)** | **0.71 (0.70-0.71)** |
| Arrhythmia | 1.71 (1.24-2.36) | 1.03 (0.83-1.31) | 1.10 (0.89-1.39) | **1.42 (1.39-1.45)** |
| Benign prostatic hyperplasia | - | **0.28 (0.23-0.34)** | - | - |
| Bipolar disorder | 1.22 (1.05-1.42) | **0.055 (0.049-0.063)** | **0.076 (0.070-0.083)** | **0.87 (0.83-0.91)** |
| Chronic air disease | **0.71 (0.66-0.77)** | **0.69 (0.65-0.75)** | **0.64 (0.61-0.67)** | **0.75 (0.75-0.76)** |
| Congestive heart failure | **1.53 (1.38-1.68)** | **0.72 (0.67-0.78)** | **0.82 (0.77-0.87)** | **1.29 (1.28-1.30)** |
| Dementia | 1.97 (0.46-8.47) | 1.63 (0.70-5.27) | 2.79 (1.19-9.00) | **0.88 (0.84-0.93)** |
| Depression | **0.80 (0.71-0.90)** | **0.027 (0.025-0.030)** | **0.032 (0.030-0.034)** | **0.62 (0.61-0.62)** |
| Diabetes | 1.03 (0.91-1.16) | **0.73 (0.66-0.81)** | **0.62 (0.59-0.67)** | **1.21 (1.19-1.22)** |
| Epilepsy | 1.01 (0.90-1.14) | **0.28 (0.26-0.32)** | **0.28 (0.26-0.30)** | **0.95 (0.93-0.96)** |
| Gastro-oesophageal reflux disease | **0.81 (0.75-0.87)** | **0.59 (0.55-0.63)** | **0.52 (0.50-0.54)** | **0.85 (0.85-0.86)** |
| Glaucoma | 1.28 (0.99-1.65) | 0.70 (0.58-0.86) | 0.84 (0.72-0.99) | **1.07 (1.05-1.09)** |
| Gout | **4.84 (3.88-6.07)** | 0.98 (0.86-1.12) | 0.83 (0.70-1.00) | **4.64 (4.55-4.73)** |
| Hepatitis B | - | 3.24 (0.73-56.78) | - | **2.09 (1.83-2.40)** |
| Hepatitis C | 3.91 (0.94-19.43) | 0.73 (0.34-2.03) | 1.38 (0.53-5.56) | **2.48 (2.16-2.84)** |
| Human Immunodeficiency Virus | **98.10 (21.06-1747.38)** | **0.31 (0.22-0.44)** | 6.90 (1.56-121.20) | **4.58 (4.15-5.06)** |
| Hyperlipidaemia | **1.79 (1.63-1.97)** | **0.64 (0.60-0.69)** | **0.78 (0.74-0.83)** | **1.42 (1.41-1.44)** |
| Hypertension | 0.97 (0.86-1.10) | **0.74 (0.66-0.82)** | **0.87 (0.81-0.93)** | 0.81 (0.80-0.82) |
| Hyperthyroidism | 0.60 (0.33-1.04) | 0.67 (0.42-1.17) | 1.25 (0.99-1.61) | **0.32 (0.30-0.33)** |
| Hypothyroidism | **0.27 (0.22-0.32)** | **0.43 (0.37-0.51)** | **0.56 (0.53-0.60)** | **0.21 (0.21-0.22)** |
| Incontinence | 0.67 (0.51-0.88) | **0.44 (0.35-0.56)** | **0.49 (0.43-0.56)** | **0.61 (0.59-0.62)** |
| Inflammation/pain | **0.82 (0.76-0.89)** | **0.62 (0.59-0.67)** | **0.52 (0.50-0.54)** | 0.99 (0.98-0.99) |
| Irritable bowel syndrome | 0.64 (0.45-0.88) | 0.82 (0.62-1.12) | **0.59 (0.51-0.68)** | **0.88 (0.85-0.91)** |
| Ischaemic heart disease: Angina | **1.64 (1.32-2.04)** | 0.76 (0.65-0.89) | **0.76 (0.66-0.88)** | **1.50 (1.48-1.53)** |
| Ischaemic heart disease: Hypertension | **1.31 (1.20-1.43)** | **0.56 (0.52-0.61)** | **0.59 (0.56-0.62)** | **1.10 (1.09-1.11)** |
| Liver failure | 1.02 (0.61-1.65) | 2.00 (1.36-3.12) | **2.09 (1.62-2.77)** | **0.92 (0.90-0.95)** |
| Malignancies | **0.49 (0.37-0.63)** | 1.18 (0.94-1.52) | **0.74 (0.66-0.83)** | **0.81 (0.79-0.83)** |
| Migraine | **0.44 (0.37-0.53)** | **0.33 (0.28-0.39)** | **0.38 (0.36-0.41)** | **0.37 (0.36-0.38)** |
| Osteoporosis/Paget's disease | **0.40 (0.31-0.53)** | **0.63 (0.51-0.81)** | 0.83 (0.74-0.94) | **0.30 (0.29-0.30)** |
| Pain | **0.81 (0.76-0.87)** | **0.59 (0.55-0.62)** | **0.47 (0.46-0.49)** | **0.96 (0.95-0.97)** |
| Pancreatic insufficiency | 0.43 (0.21-0.80) | 0.83 (0.48-1.60) | **0.41 (0.32-0.54)** | **0.85 (0.80-0.90)** |
| Parkinson's disease | 0.94 (0.74-1.20) | **0.40 (0.33-0.50)** | **0.33 (0.29-0.38)** | **1.08 (1.05-1.11)** |
| Psoriasis | 1.40 (1.08-1.80) | **0.63 (0.52-0.78)** | **0.75 (0.64-0.87)** | **1.24 (1.21-1.27)** |
| Psychotic illness | 1.15 (1.04-1.26) | **0.20 (0.18-0.21)** | **0.17 (0.16-0.18)** | **1.11 (1.09-1.12)** |
| Pulmonary hypertension | - | - | 1.86 (0.42-32.71) | **0.48 (0.37-0.61)** |
| Renal disease | 0.86 (0.39-1.77) | 1.43 (0.81-2.85) | 1.30 (0.89-2.00) | 0.97 (0.93-1.02) |
| Smoking cessation | 1.15 (0.99-1.33) | 1.08 (0.96-1.23) | 0.97 (0.89-1.05) | **1.34 (1.32-1.36)** |
| Steroid-responsive disease | **0.69 (0.63-0.75)** | **0.61 (0.56-0.65)** | **0.53 (0.51-0.55)** | **0.76 (0.76-0.77)** |
| Transplant | 0.92 (0.46-1.74) | 0.65 (0.39-1.22) | **0.40 (0.29-0.57)** | **1.45 (1.34-1.57)** |
| Tuberculosis | 1.00 (0.05-10.77) | 0.89 (0.20-15.70) | 1.01 (0.32-6.10) | 1.23 (0.98-1.53) |

Abbreviations: 10PCT, 10% Sample of the Australian PBS Claims Database; AGDS, Australian Genetics of Depression Study; OR (95% CI), Adjusted odds ratio (95% confidence interval). Odds ratios were adjusted for participant age and those highlighted in bold met the Bonferroni threshold for multiple testing (0.05/164=0.000305).

Compared to the general population, AGDS participants had significantly higher odds of having inferred comorbidities that are known to be more prevalent among people living with depression, including other mental health conditions, pain, diabetes and cardiovascular conditions (Table 2). Notably, the highest risk of the cardiovascular conditions (hypertension related to ischaemic heart disease, congestive heart failure, hyperlipidaemia, hypertension) in the AGDS was observed among male participants, while the highest risk of all mental health conditions, except for alcohol dependency, was observed among female participants.

### Impact of comorbidities on health care costs in the AGDS

Cost data in the linked MBS and PBS records of 11,186 female and 3,836 male AGDS participants were explored to examine the impact of multimorbidity on health care costs, both with respect to out-of-pocket expenses for the individual and costs to the Medicare health system. The median annual Medicare benefits paid (MBS plus PBS costs) for male (median, $1503.21; interquartile range [IQR], $789.72-2834.44) and female (median, $1576.18; IQR, $918.98-2705.04) participants were similar, as were the median annual out-of-pocket expenses for male (median, $613.77; IQR, $311.25-1202.68) and female (median, $624.52; IQR, $313.97-1424.03) participants. On average, however, male participants were dispensed significantly more prescriptions than females (27.2 vs 22.9; *p* < 0.001) but accessed significantly fewer medical services (26.7 vs 30.1; *p* < 0.001) per year.

Participant remoteness also significantly impacted the annual number of services accessed (*p* = 0.0045), where participants living in metropolitan areas accessed more services (*n* = 10512, *x̅* = 29.7, *s* = 24.2) compared to those in regional or rural areas (*n* = 4106, *x̅* = 28.2, *s* = 21.2) and remote or very remote areas (*n* = 187, *x̅* = 25.6, *s* = 17.6). In addition, the annual number of services accessed was positively correlated with participant age (*r_s_*(15022) = 0.141, 95% BCa CI [0.126, 0.157], *p* < 0.001) and severity of illness indicators such as the number of DSM5 MDD symptoms (*r_s_*(12895) = 0.102, 95% BCa CI [0.084, 0.120], *p* < 0.001) and depressive episodes (*r_s_*(13119) = 0.130, 95% BCa CI [0.113, 0.147], *p* < 0.001). Similarly, the annual number of prescriptions dispensed was significantly correlated (*p* < 0.001) with participant age (*r_s_*(15022) = 0.438, 95% BCa CI [0.425, 0.452]), number of DSM5 MDD symptoms (*r_s_*(12895) = 0.036, 95% BCa CI [0.019, 0.053]) and number of depressive episodes (*r_s_*(13119) = 0.167, 95% BCa CI [0.150, 0.183]). However, participants living in metropolitan areas were dispensed fewer prescriptions (*x̅* = 22.8, *s* = 22.2) compared to those in regional or rural areas (*x̅* = 26.7, *s* = 24.2) and remote or very remote areas (*x̅* = 26.1, *s* = 26.2).

Spearman’s rank-order correlations show significant positive relationships between the Rx-Risk score and the annual patient out-of-pocket costs (*r_s_*(15022) = 0.380, 95% BCa CI [0.366, 0.395], *p* < 0.001) and Medicare benefits paid (*r_s_*(15022) = 0.630, 95% BCa CI [0.619, 0.640], *p* < 0.001) costs. After excluding the depression comorbidity category when calculating the Rx-Risk score, the annual Medicare benefits paid per participant with no comorbidities (median, $733.10; IQR, $451.77-1170.82) was less for participants with one (median, $993.62; IQR, $578.13-1394.68) or two (median $1089.00; IQR, $708.80-1680.42) comorbidities. Similarly, median annual out-of-pocket expenses were lowest for participants with no comorbidities (median, $332.56; IQR, $167.11-584.94) compared to those with one (median, $420.71; IQR, $236.61-733.38) or two (median, $514.91; IQR, $264.95-889.47). Compared to participants with one or two comorbidities, participants with no comorbidities were also dispensed fewer prescriptions (8.6 vs 10.8 and 13.5 respectively) and accessed fewer health services (15.1 vs 18.0 and 20.7 respectively) on average per year. The rise in annual out-of-pocket costs and Medicare benefits paid as the participant’s number of comorbidities increased is summarised in Table 3 and illustrated in Supplemental Figure S6.

**Table 3.** Annual combined PBS and MBS benefits paid and patient out-of-pocket costs (AUD) by Rx-Risk score of Australian Genetics of Depression Study participants.

| Rx-Risk score<sup>a</sup> | Participants (n) | Medicare benefits paid |  |  | Patient out-of-pocket costs |  |  |
| --- | --- | --- | --- | --- | --- | --- | --- |
|  |  | Median (IQR) | Mean (95% CI) | SD | Median (IQR) | Mean (95% CI) | SD |
| 0 | 1428 | \$733.10<br>(\$451.78, \$1,170.82) | 1,024.90<br>(\$951.06, \$1,098.74) | \$1,422.37 | \$332.56<br>(\$167.11, \$584.94) | \$477.10<br>(\$448.63, \$505.58) | \$548.48 |
| 1 | 2006 | \$933.62<br>(\$578.13, \$1,394.68) | 1,193.71<br>(\$1,133.11, \$1,254.30) | \$1,786.68 | \$420.71<br>(\$236.61, \$733.38) | \$579.37<br>(\$554.87, \$603.88) | \$559.59 |
| 2-3 | 4449 | \$1,213.95<br>(\$778.99, \$1,867.65) | 1,563.23<br>(\$1,519.89, \$1,606.58) | \$1,474.73 | \$562.17<br>(\$289.83, \$1,005.92) | \$767.67<br>(\$746.32, \$789.03) | \$726.54 |
| 4-6 | 4536 | \$1,977.64<br>(\$1,302.67, \$2,959.39) | 2,449.53<br>(\$2,392.32, \$2,506.74) | \$1,965.33 | \$800.08<br>(\$389.00, \$1,449.14) | \$1,078.55<br>(\$1,050.00, \$1,107.09) | \$980.66 |
| 7-10 | 2220 | \$3,502.36<br>(\$2,357.51, \$5,219.78) | 4,155.78<br>(\$4,042.66, \$4,268.91) | \$2,718.03 | \$1,194.21<br>(\$473.41, \$2,223.90) | \$1,573.89<br>(\$1,513.85, \$1,633.93) | \$1,442.65 |
| 11-14 | 360 | \$5,814.94<br>(\$4,318.40, \$8,100.28) | \$6,536.85<br>(\$6,176.85, \$6,896.86) | \$3,473.28 | \$1,339.79<br>(\$491.32, \$2,947.47) | \$1,971.65<br>(\$1,784.34, \$2,158.95) | \$1,807.11 |
| 15-18 | 23 | \$10,815.10<br>(\$7,854.15, \$13,909.61) | \$12,378.76<br>(\$9,081.07, \$15,676.46) | \$7,625.92 | \$2,456.38<br>(\$1,275.40, \$4,600.65) | \$3,151.79<br>(\$1,937.41, \$4,366.17) | \$2,808.25 |

Abbreviations: 95% CI, 95% confidence intervals; IQR, interquartile range; n, number of participants; SD, standard deviation. <sup>a</sup>Defined as the number of different comorbidity categories after excluding depression mapped by prescriptions dispensed between 01/07/2013 and 31/12/2017.

## 4. Discussion

Using Medicare claims data, we measured chronic disease status for 46 categories of comorbidity, including both psychiatric and physical disorders, in a depressed cohort and an unselected Australian population sample, and explored the impact of chronic disease burden on health care costs to the individual and the government. Given that we did not have access to hospital diagnostic records in either the AGDS or 10PCT samples, the validated medication-based Rx-Risk Comorbidity Index is a useful tool to infer a participant’s comorbidities in the Australian setting.

Our findings extend previous research by demonstrating a higher burden of inferred physical and mental health comorbidities among participants in a retrospective cohort study of Australians living with severe depression (AGDS; mean [SD]: 4.6 [2.9]) compared to an unselected Australian population sample (10PCT; mean [SD]: 3.0 [3.0]). As expected, depression was the most inferred comorbidity among 90.7% of AGDS participants. In comparison, 23.1% of individuals in the 10PCT sample had medications dispensed that mapped to the depression comorbidity (specifically, non-selective monoamine reuptake inhibitors, selective serotonin reuptake inhibitors, monoamine oxidase A inhibitors, non-selective monoamine oxidase inhibitors, non-selective and other antidepressants in the NO6AX ATC category).

Given that the lifetime prevalence of major depression was 10.3% in South Australia in 2008 [19] and the lifetime and 12-month prevalence rates for a depressive episode were 11.7% and 4.9% respectively among Australians aged 16-85 years in the National Study of Mental Health and Wellbeing (NSMWH) 2020-2022 survey [20], this may indicate that the depression category is over-estimated by the Rx-Risk Comorbidity Index. A potential reason for this over-estimation is that antidepressants have more than one indication. For example, while antidepressants are commonly used as first-line treatments for anxiety disorders [21], the Rx-Risk Comorbidity Index model only maps benzodiazepine derivative drugs and buspirone to the anxiety comorbidity category. Subsequently, this may be the reason that the 10.8% prevalence rate of inferred anxiety in the unselected Australian population sample (10PCT) was much lower than expected given that the 12-month prevalence rate of anxiety disorders observed in the NSMWH 2020-2022 survey was 17.2% [20].

With respect to physical comorbidities among individuals living with depression, male AGDS participants were at highest risk of HIV, gout, hepatitis C, and antiplatelet use and lowest risk for hypothyroidism and migraine compared to female AGDS participants. Male AGDS participants were also at significantly higher risk for ischaemic heart disease and cardiovascular disease traits (congestive heart failure, hyperlipidaemia).

Compared to the unselected Australian population sample, AGDS participants were at higher risk of the other mental health conditions among the 46 inferred comorbidity categories; AGDS participants were at >10-fold increased risk of bipolar disorder, >5-fold increased risk of alcohol dependency and psychotic illness, and ∼4-fold increased risk of anxiety. High rates of comorbidity between depression and other mental health conditions, in particular anxiety, has long been established [22]. AGDS male participants were also at increased risk for 29 out of 37 inferred physical health conditions with 21 surviving correction for multiple testing. AGDS males had more than 3-fold higher risk of being classified as having migraines, HIV, and epilepsy compared to the Australian population and this is in line with previous research [23–26]. Female AGDS participants also had more than 3-fold higher risk for epilepsy and had >2-times higher risk for migraines and transplant. However, it must be noted that our findings do not take in to account the direction of causality which may bias the prevalence of each physical health condition. For example, depression is the most frequent comorbid psychiatric disorder in epilepsy and this relationship is generally interpreted as depression arising due to the experience of living with epilepsy [27].

Overall, our findings align with those from a 2023 UK Biobank study [22] that investigated the association between depression and 77 health conditions requiring hospitalization. In that study, following adjustment for age, sex, ethnic origin, education, smoking status, alcohol consumption, and physical activity, and confirmation in a Finnish cohort, participants living with severe/moderately severe depression had at least 1.5-times higher risk of 27 conditions across multiple organ systems including mental health and behavioural disorders (hazard ratio [HR], 5.97; 95% CI, 3.97-8.99), Parkinson’s disease (HR, 16.47; 95% CI, 6.77-40.06), ischemic heart disease (HR, 1.76; 95% CI, 1.36-2.29), heart failure (HR, 4.38; 95% CI, 2.66-7.23), diabetes (HR, 5.15; 95% CI, 2.52-10.50), chronic obstructive bronchitis (HR, 4.11; 95% CI, 2.56-6.60), headaches (HR, 3.67; 95% CI, 2.16-6.25), gout (HR, 6.80; 95% CI, 2.32-19.93), and back pain (HR, 3.99; 95% CI, 2.96-5.38). Severely/moderately severely depressed individuals were also at higher risk of hypertension (HR=1.47; 95% CI, 0.60-3.62) but the association was not significant. While depression was associated with increased risk for all circulatory system diseases in the UK Biobank study, a significant increased risk for angina was only observed for female participants when comparing the AGDS and 10PCT samples. Female AGDS participants had higher risk for all cardiovascular-related comorbidity categories than females in the 10PCT sample which agrees with prior research that found that depression in women was a significant risk factor for the development of cardiovascular disease [28, 29].

Finally, we evaluated annual health care costs in the 4.5-year window for all AGDS participants. These health care costs were driven by increased frequency in both PBS (dispensed prescriptions) and MBS (medical service utilization) items. Both the median annual out-of-pocket costs and Medicare benefits paid both rose more than 65% when comparing participants with no inferred comorbidities to those with 2-3 comorbidities. Notably, the increase in Medicare benefits paid was two-folder higher than that of out-of-pocket costs among participants with 11-13 and 15-18 comorbidities. This means that the Australian government is making substantially higher payments for individuals living with depression who are also experiencing a high burden of comorbidities. The association between multimorbidity and increased healthcare costs and utilization was also reported in systematic reviews of 17 UK studies [30]. Critically, a UK study of the impact of comorbidity on primary care costs found that depression was the main cost-increasing comorbidity when it co-occurred with a range of conditions across all ages [31].

### 4.1 Strengths and Limitations

A major strength of our study is the use of individual-level Medicare claims data in a depressed cohort as well as PBS data in a large, unselected population representative sample. This objective real-world data does not rely on self-report information collected from participants. Notably, individuals with lived experience of depression were not excluded from the population representative sample. However, given the high proportion of AGDS participants who live with recurrent severe depression, our findings may not generalize to all Australians with lived experience of depression.

The prescription-based Rx-Risk Comorbidity Index also has potential limitations with respect to the accuracy of the rates of each inferred comorbidity. First, we have potentially shown that a participant’s age, remoteness and severity of illness may impact on their ability to access medical services, as well as their frequency of access, and the pattern of prescriptions that the participant’s health care provider prescribed. All these factors may result in bias in the prescription dispensing records and the inference of comorbidities and as such will lower the prognostic accuracy of the Rx-Risk Comorbidity Index. Second, the use of dispensed medications, which may have a main indication but could be used to treat more than one disease, as a proxy for inferring comorbidities may result in misclassification of some chronic conditions. Consequently, prevalences of one or more of the Rx-Risk comorbidity categories may be under- or over-estimated. Third, the PBS records that are maintained by the *Services Australia* do not contain records of prescriptions dispensed in hospital settings, private prescriptions (e.g. where a medication is only listed on the PBS for individuals over a certain age), or purchases of medications over the counter at pharmacies where a prescription is not required such as commonly used anti-histamines. Therefore, the prescription dispensing data may not have captured all medications used by individuals in the AGDS and 10PCT samples in the 4.5-year window.

## 5. Conclusions

We found a higher burden of physical and mental health conditions among people living with depression, and that burden was positively associated with age, BMI, and number of depressive episodes. We also observed a strong economic burden of multimorbidity where health care costs to the participant and the Medicare health system increased along with their number of inferred comorbidities. These findings suggest, Australians living with depression and high rates of multimorbidity may experience increased out-of-pocket health care costs which may limit their ability to access services. Our research findings improve our understanding of the causes of excess morbidity among people with lived experience of depression, which is a public health priority [8].

## Acknowledgments

We thank all the participants for giving their time to contribute to this study. We wish to thank all the people who helped in the conception, implementation, beta testing, media campaign and data cleaning. We would specifically like to acknowledge Ken Kendler, Patrick Sullivan, Andrew McIntosh and Cathryn Lewis for input on the questionnaire; Lorelle Nunn, Mary Ferguson, Lucy Winkler and Natalie Garden for data and sample collection; Jonathan Davies, Luke Lowrey and Valeriano Antonini for support with IT aspects; Vera Morgan and Ken Kirkby for help with the media campaign.

## CRediT authorship contribution statement

**Penelope A Lind**: Writing – original draft, Data curation; Methodology, Formal analysis, Conceptualization. **Ian B. Hickie**: Writing – review & editing; Funding acquisition. **Enda M Byrne**: Writing – review & editing; Funding acquisition. **Nicholas G. Martin**: Writing – review & editing, Data curation; Funding acquisition. **Sarah E Medland**: Writing – review & editing; Data curation; Funding acquisition.

## Supplemental data

Supplemental data to this article can be found online.

## Data availability

Anonymised survey data may be shared for collaborative projects, subject to a data transfer agreement and governance and ethics approval. Pharmaceutical Benefits Scheme and Medicare Benefits Schedule data cannot be shared due to legal restrictions.

