## Supplemental for "Burden of health morbidities and associated health care costs in the Australian Genetics of Depression Study using the medication-based Rx-Risk Comorbidity Index"

**Contents**

**Table S1.** Prevalence of each inferred comorbidity category by sample and sex. .................... 2

**Figure S1.** Distribution of participant age by sex in the Australian Genetics of Depression Study (AGDS) and 10% Sample of the Australian PBS Claims Database (10PCT) samples... 4

**Table S1.** Prevalence of each inferred comorbidity category by sample and sex.

| Comorbidity Category | AGDS | | | 10PCT | | |
|---|---|---|---|---|---|---|
| | Male ($n$ = 4196) | Female ($n$ = 11694) | Total ($n$ = 15890) | Male ($n$ = 775683) | Female ($n$ = 860795) | Total ($n$ = 1636478) |
| Alcohol dependency | 2.6% | 1.7% | 1.9% | 0.50% | 0.29% | 0.4% |
| Allergies | 0.0% | 0.060% | 0.0% | 0.022% | 0.039% | 0.0% |
| Anticoagulants | 7.6% | 6.7% | 7.0% | 6.9% | 6.8% | 6.9% |
| Antiplatelets | 6.7% | 2.1% | 3.3% | 9.0% | 6.6% | 7.7% |
| Anxiety | **29.9%** | **33.2%** | **32.3%** | 9.1% | **12.3%** | **10.8%** |
| Arrhythmia | 1.8% | 0.69% | 1.0% | 2.7% | 2.0% | 2.4% |
| Benign prostatic hyperplasia | 2.6% | 0.026% | 0.7% | 1.2% | 0.011% | 0.6% |
| Bipolar disorder | 6.6% | 5.6% | 5.8% | 0.39% | 0.44% | 0.4% |
| Chronic air disease | **24.3%** | **29.6%** | **28.2%** | **18.4%** | **23.0%** | **20.8%** |
| Congestive heart failure | **23.6%** | **12.9%** | **15.7%** | **20.9%** | **17.8%** | **19.3%** |
| Dementia | 0.095% | 0.034% | 0.1% | 0.39% | 0.49% | 0.4% |
| Depression | **89.2%** | **91.3%** | **90.7%** | **18.7%** | **27.1%** | **23.1%** |
| Diabetes | **10.1%** | 8.9% | 9.2% | 8.6% | 7.3% | 7.9% |
| Epilepsy | **10.3%** | 10.0% | **10.1%** | 3.2% | 3.4% | 3.3% |
| Gastrooesophageal reflux disease | **35.9%** | **36.6%** | **36.4%** | **26.1%** | **28.7%** | **27.5%** |
| Glaucoma | 2.6% | 1.3% | 1.6% | 2.5% | 2.4% | 2.5% |
| Gout | 6.0% | 1.0% | 2.3% | 6.6% | 1.6% | 4.0% |
| Hepatitis B | 0.024% | 0.0% | 0.0% | 0.078% | 0.037% | 0.1% |
| Hepatitis C | 0.12% | 0.026% | 0.1% | 0.087% | 0.035% | 0.1% |
| HIV | 0.83% | 0.009% | 0.2% | 0.26% | 0.057% | 0.2% |
| Hyperkalaemia | 0.0% | 0.0% | 0.0% | 0.000% | 0.000% | 0.0% |
| Hyperlipidaemia | **28.6%** | **14.2%** | **18.0%** | **23.6%** | **19.2%** | **21.3%** |
| Hypertension | **10.2%** | 7.7% | 8.4% | 9.1% | **10.7%** | 9.9% |
| Hyperthyroidism | 0.36% | 0.56% | 0.5% | 0.25% | 0.79% | 0.5% |
| Hypothyroidism | 3.7% | **10.3%** | 8.5% | 1.8% | 7.7% | 4.9% |
| Incontinence | 1.7% | 2.0% | 1.9% | 1.0% | 1.6% | 1.3% |
| Inflammation/pain | **39.0%** | **40.8%** | **40.3%** | **28.8%** | **29.0%** | **28.9%** |
| Irritable bowel syndrome | 1.1% | 1.6% | 1.4% | 0.90% | 1.0% | 1.0% |
| Ischaemic heart disease: Angina | 3.9% | 1.7% | 2.3% | 4.1% | 2.9% | 3.5% |
| Ischaemic heart disease: Hypertension | **23.0%** | **15.6%** | **17.5%** | **17.2%** | **16.0%** | **16.6%** |
| Liver failure | 0.55% | 0.47% | 0.5% | 1.4% | 1.5% | 1.4% |
| Malignancies | 1.6% | 2.6% | 2.4% | 2.1% | 2.6% | 2.4% |
| Malnutrition | 0.0% | 0.0% | 0.0% | 0.012% | 0.010% | 0.0% |
| Migraine | 3.6% | 8.2% | 7.0% | 1.2% | 3.3% | 2.3% |
| Osteoporosis/Pagets | 1.8% | 2.5% | 2.3% | 1.8% | 5.3% | 3.6% |
| Pain | **48.1%** | **52.1%** | **51.0%** | **35.3%** | **36.2%** | **35.8%** |
| Pancreatic insufficiency | 0.26% | 0.51% | 0.4% | 0.23% | 0.28% | 0.3% |
| Parkinsons disease | 2.3% | 2.0% | 2.1% | 1.2% | 1.1% | 1.1% |
| Psoriasis | 2.3% | 1.5% | 1.7% | 1.5% | 1.2% | 1.3% |
| Psychotic illness | **17.9%** | **16.3%** | **16.7%** | 4.1% | 3.7% | 3.9% |
| Pulmonary hypertension | 0.0% | 0.009% | 0.0% | 0.011% | 0.024% | 0.0% |
| Renal disease | 0.24% | 0.21% | 0.2% | 0.43% | 0.45% | 0.4% |

| | | | | | | |
|---|---|---|---|---|---|---|
| Smoking cessation | 6.6% | 5.7% | 5.9% | 7.1% | 5.4% | 6.2% |
| Steroid-responsive disease | **21.4%** | **26.5%** | **25.2%** | **14.6%** | **18.0%** | **16.4%** |
| Transplant | 0.29% | 0.32% | 0.3% | 0.19% | 0.129% | 0.2% |
| Tuberculosis | 0.024% | 0.017% | 0.0% | 0.021% | 0.017% | 0.0% |

Abbreviations: 10PCT, 10% Sample of the Australian PBS Claims Database; AGDS, Australian Genetics of Depression Study. Prevalence rates greater than 10% are highlighted in bold.

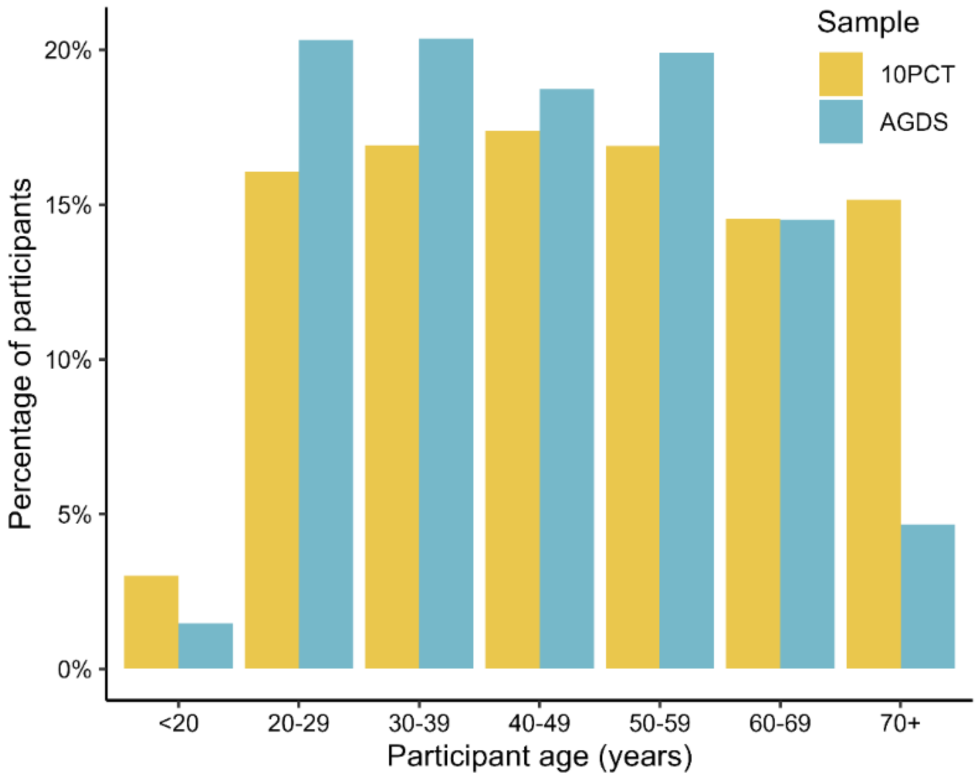

**Figure S1.** Distribution of participant age by sex in the Australian Genetics of Depression Study (AGDS) and 10% Sample of the Australian PBS Claims Database (10PCT) samples.

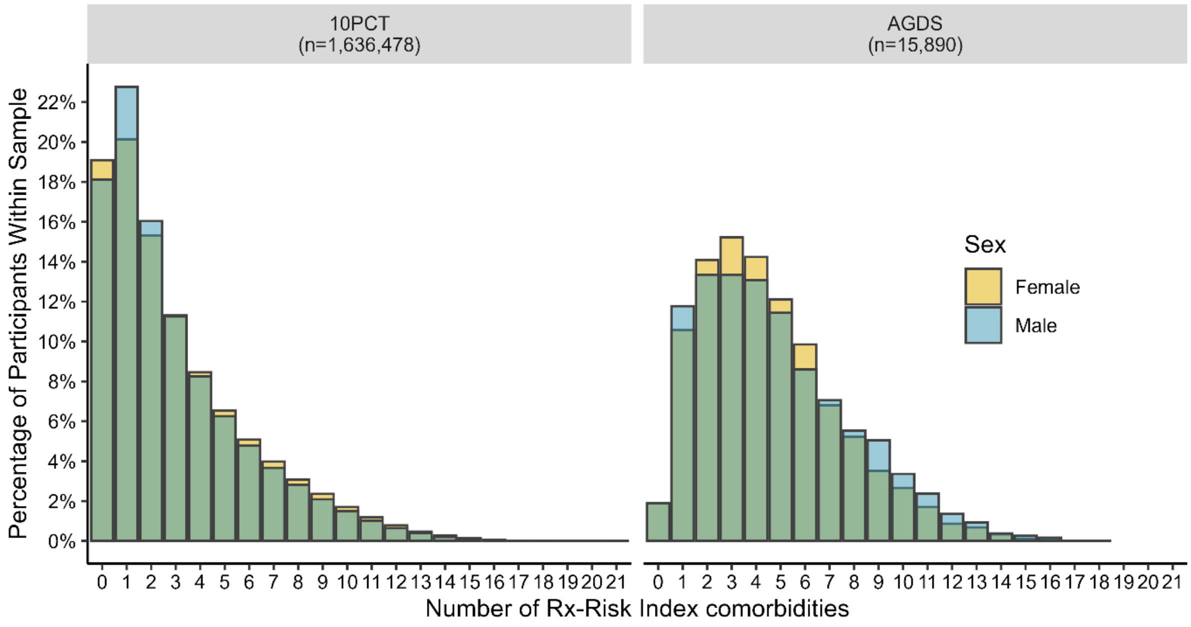

**Figure S2.** Distribution of the number of Rx-Risk Index comorbidities[a] by sex in the Australian Genetics of Depression Study (AGDS) and 10% Sample of the Australian PBS Claims Database (10PCT) samples. Where the percentage of male (grey) and female (yellow) participants overlap the bar plot is green.

[a]Defined as the number of different comorbidity categories mapped by prescriptions dispensed between 01/07/2013 and 31/12/2017, possible range 0-46.

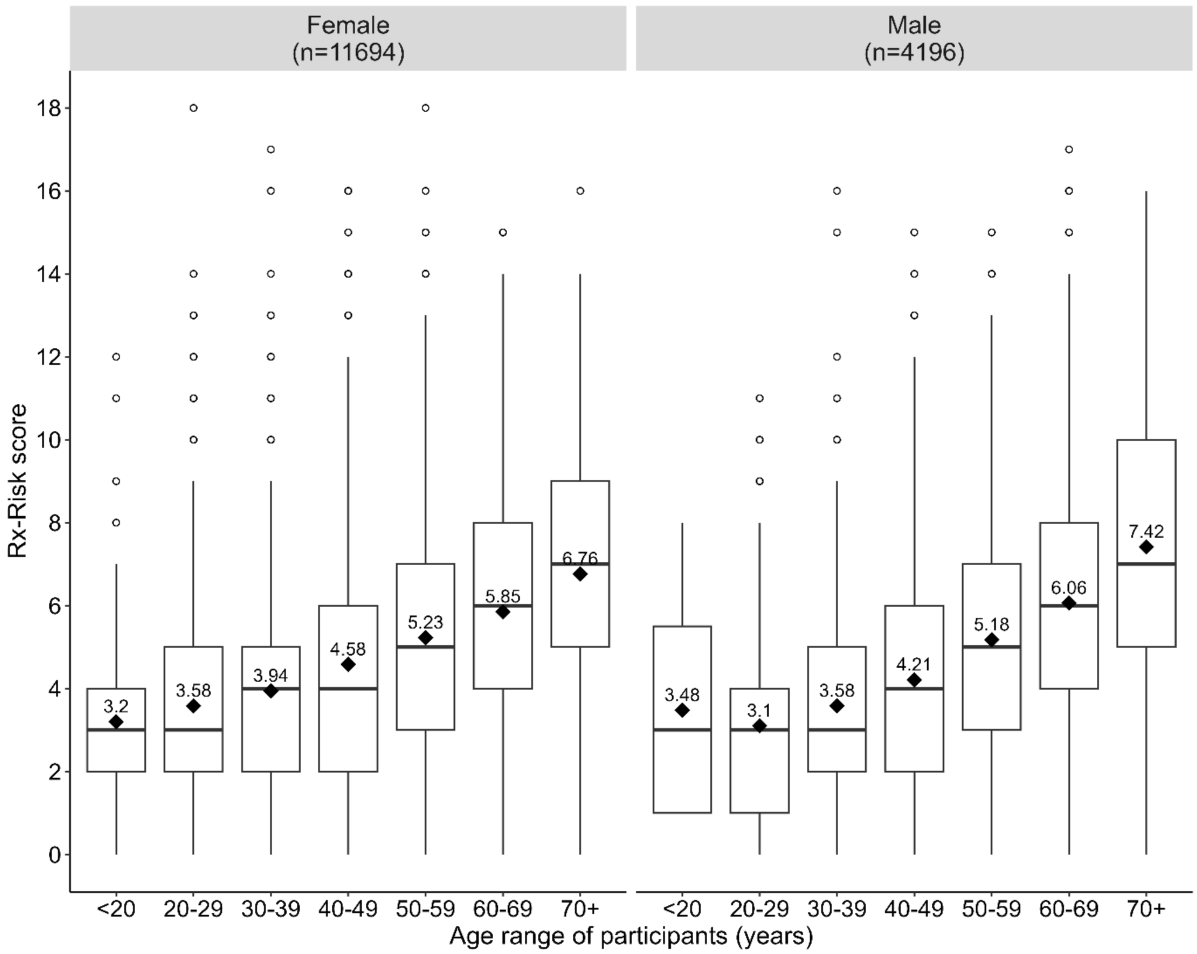

**Figure S3**. Box plots of Rx-Risk scores[a] by sex and age range of Australian Genetics of Depression Study participants. Black diamonds and numbers represent the mean Rx-Risk score for each age range. The black bar represents the median.

[a]Defined as the number of different comorbidity categories mapped by prescriptions dispensed between 01/07/2013 and 31/12/2017, possible range 0-46.

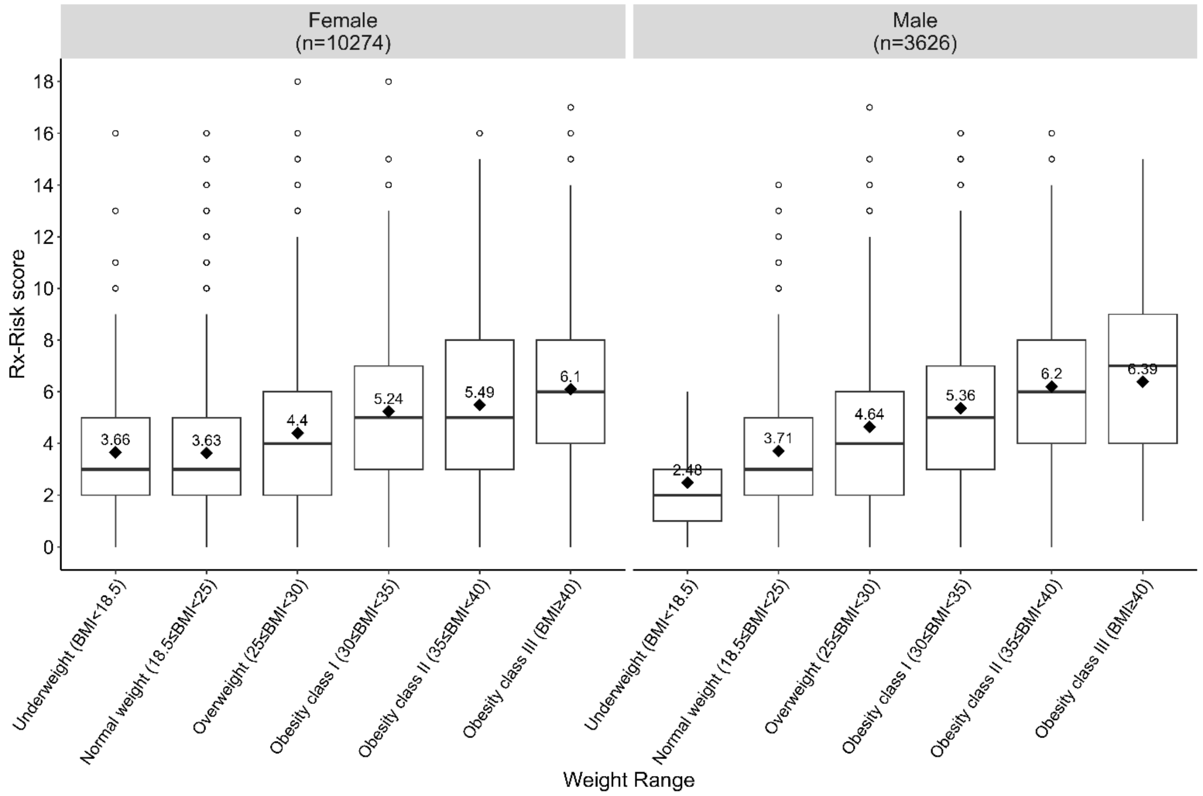

**Figure S4**. Box plots of Rx-Risk scores[a] by sex and self-reported obesity level by Australian Genetics of Depression Study participants. Black diamonds and numbers represent the mean Rx-Risk score for each obesity class. The black bar represents the median.

[a]Defined as the number of different comorbidity categories mapped by prescriptions dispensed between 01/07/2013 and 31/12/2017, possible range 0-46.

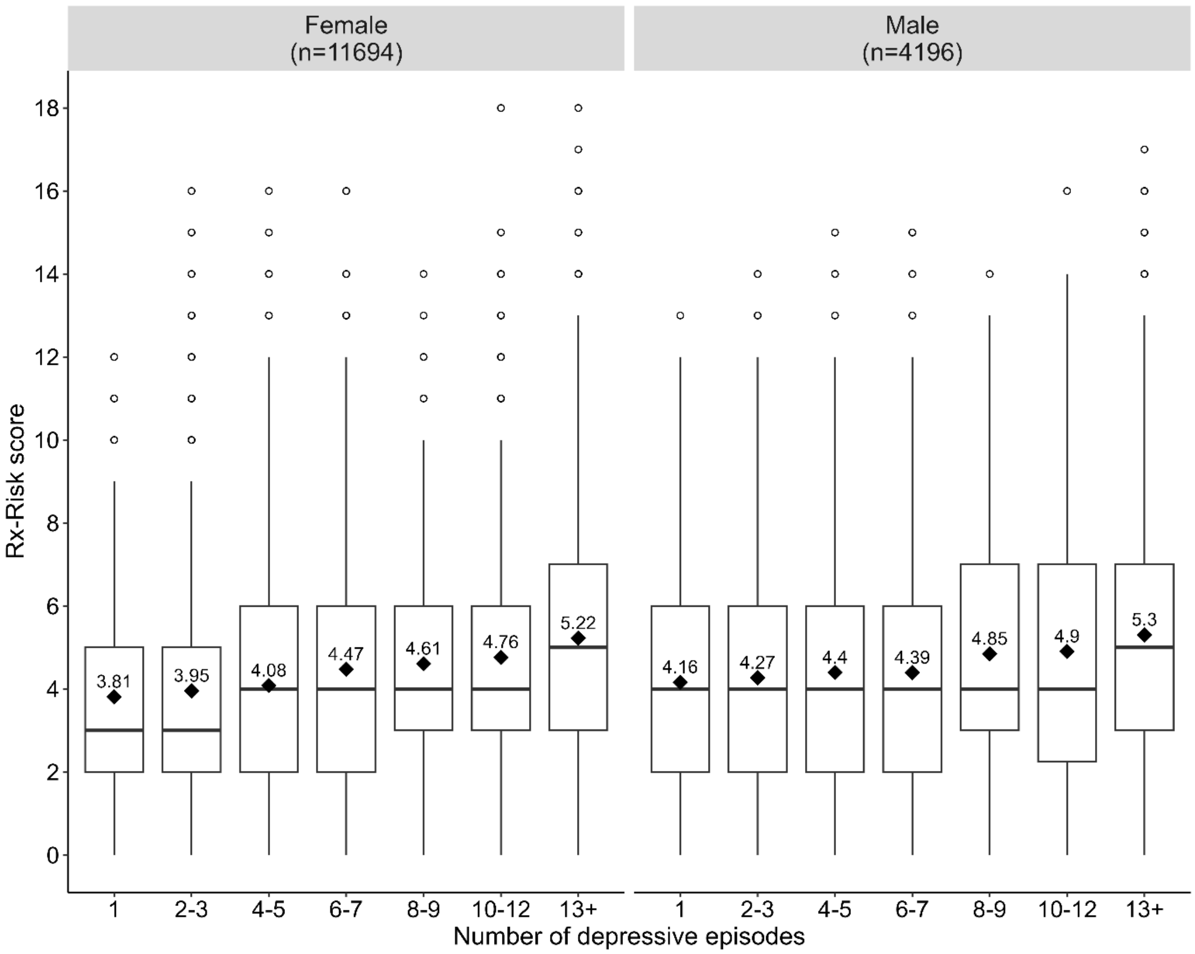

**Figure S5**. Box plots of Rx-Risk scores[a] by sex and the self-reported number of depressive episodes experienced by Australian Genetics of Depression Study participants. Black diamonds and numbers represent the mean Rx-Risk score for each depressive episode range. The black bar represents the median.

[a]Defined as the number of different comorbidity categories mapped by prescriptions dispensed between 01/07/2013 and 31/12/2017, possible range 0-46.

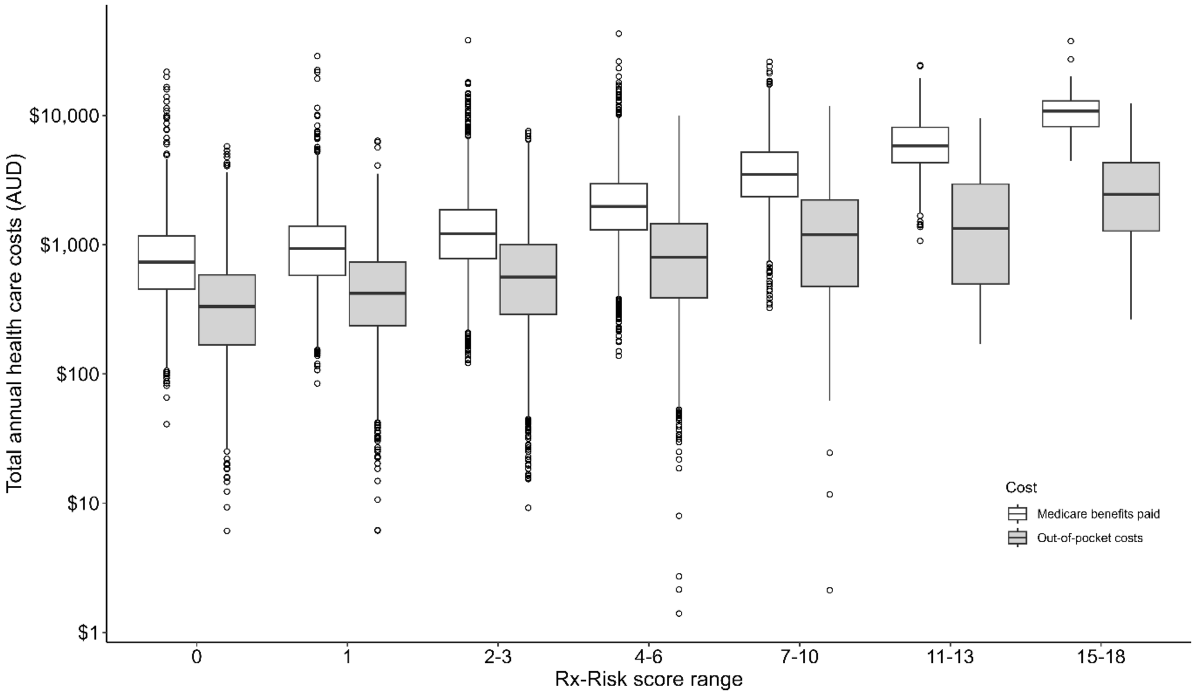

**Figure S6**. Box plots of annual out-of-pocket costs and Medicare benefits paid by Rx-Risk score[a] of Australian Genetics of Depression Study participants. The black bar represents the median.

[a]Defined as the number of different comorbidity categories after excluding depression mapped by prescriptions dispensed between 01/07/2013 and 31/12/2017, possible range 0-46.
